# Efficacy of external diaphragm pacing combined with high-flow oxygen via tracheostomy for weaning in patients requiring prolonged mechanical ventilation: a study protocol for a randomized controlled trial

**DOI:** 10.64898/2026.03.07.26347833

**Authors:** Lei Yang, Kebin Zheng, Na Li, Kongting Xing, Jing Zhang

## Abstract

**Background:** Prolonged mechanical ventilation (PMV)—commonly defined as requiring at least 6 h per day of invasive mechanical ventilation for more than 21 consecutive days—is a major challenge in critical care, associated with prolonged weaning, high ventilator-associated complication rates, and substantial morbidity and mortality. A key driver of poor outcomes is ventilator-induced diaphragmatic dysfunction (VIDD), characterized by rapid diaphragmatic atrophy and functional decline. External diaphragm pacing (EDP), a non-invasive phrenic nerve stimulation technique, may enhance diaphragmatic contractility, while high-flow oxygen via tracheostomy (HFT) can optimize oxygenation, reduce respiratory workload, and improve comfort during weaning. However, no randomized controlled trial (RCT) has evaluated the combined effect of EDP and HFT on weaning outcomes in patients requiring PMV.

**Methods:** This is a single-center, randomized, controlled, open-label, trial conducted in the intensive care unit (ICU) of a rehabilitation hospital in Ningbo, China. We will enroll 60 adult tracheostomized patients with PMV and randomize them 1:1 to an experimental group (EDP + HFT) or a control group (standard care alone). The primary endpoint is time (days) from randomization to successful 24-h separation from invasive mechanical ventilation by day 28, without reconnection to the ventilator within the subsequent 48 h. Randomization will use a computer-generated permuted block design, with allocation concealed in sequentially numbered, opaque, sealed envelopes. Treating clinicians and patients will not be blinded, but diaphragm ultrasound assessors and statisticians will remain blinded to group allocation. Recruitment is planned from 1 November 2025 to 30 April 2026.

**Discussion:** This RCT will evaluate the efficacy and safety of combining non-invasive EDP with HFT to assist weaning in tracheostomized patients requiring PMV. If EDP + HFT is associated with reduced time to ventilator liberation and improved diaphragm function, it could serve as an accessible strategy to mitigate VIDD and improve weaning outcomes.

**Trial registration:** Chinese Clinical Trial Registry (ChiCTR): ChiCTR2500098295 (registered on 5 March 2025).

**Administrative information:** Note: The numbers in curly brackets refer to SPIRIT checklist item numbers. The order is modified to optimize flow (see SPIRIT 2013 statement).

| Item | Details |
| --- | --- |
| Title {1} | Efficacy of external diaphragm pacing combined with high-flow oxygen via tracheostomy for weaning in patients requiring prolonged mechanical ventilation: a study protocol for a randomized controlled trial |
| Trial registration {2a and 2b} | Chinese Clinical Trial Registry (ChiCTR): ChiCTR2500098295 (registered 5 March 2025). The registry complies with the WHO Trial Registration Data Set. The anticipated recruitment period is from 1 November 2025 to 30 April 2026. |
| Protocol version {3} | Version 1.1, approved on 1 March 2025. |
| Funding {4} | Supported by the Medical Science and Technology Project of Zhejiang Province (Grant No. 2024KY360). The funder has no role in study design, data |
|  | collection/analysis/interpretation, manuscript writing, or submission decisions. |
| Author details {5a} | See Title Page |
| Trial sponsor {5b} | Ningbo Zhenhai Longsai Hospital, Ningbo Zhenhai 315299, Zhejiang Province, China. Contact: Medical Research Office, |
| Role of sponsor {5c} | The sponsor has no authority over study design, data management, analysis, or publication decisions. |
| Trial management group {5d} | The Trial Management Group (TMG) includes the principal investigator (intensivist), a co-investigator from the ultrasound department, the research nurse, and the biostatistician. The TMG meets at least monthly to review recruitment, protocol adherence, data quality, and safety reports and to address operational issues. |

## Introduction

### Background and rationale {6a}

Prolonged mechanical ventilation (PMV), typically defined as requiring at least 6 h per day of mechanical ventilation for more than 21 consecutive days [1, 2], is a critical intervention for patients with respiratory failure. PMV is frequently associated with prolonged weaning, defined by the International Task Force as failing at least three weaning attempts or requiring more than 7 days of weaning after the first spontaneous breathing trial [3–5]. With advancements in intensive care, the number of patients surviving acute respiratory failure and requiring PMV has risen significantly [6–9]. These patients consume substantial healthcare resources and face high short-and long-term mortality rates [10–13].

PMV and prolonged weaning are linked to increased ventilator-associated complications, creating a vicious cycle of morbidity and mortality. A key contributor is ventilator-induced diaphragmatic dysfunction (VIDD), characterized by rapid diaphragmatic atrophy and functional decline due to mechanical ventilation [14, 15]. VIDD delays liberation from mechanical ventilation, prolongs weaning, and increases dependency on ventilatory support, ultimately impairing patients’ quality of life [16–18].

To address these issues, strategies such as diaphragm pacing have been explored. Phrenic nerve stimulation has demonstrated potential in alleviating VIDD and enhancing diaphragm training, thereby aiding weaning [19]. Traditional methods (e.g., surgically implantable devices) are effective but invasive, technically demanding, and costly [20–28]. In contrast, non-invasive external diaphragm pacing (EDP) delivers targeted electrical stimuli through the skin, offering a less invasive alternative to enhance diaphragm contractility [29–31].

High-flow oxygen via tracheostomy (HFT) is another innovative approach for PMV weaning [32–36]. HFT improves oxygenation, maintains lung volumes, reduces work of breathing, generates positive end-expiratory pressure, and facilitates CO₂ washout. Preliminary data in tracheostomized populations suggest that HFT may reduce re-intubation risk, stabilize oxygenation, and be well tolerated compared with non-invasive ventilation [33, 36].

Although both EDP and HFT are non-invasive, they are not entirely risk-free. Reported adverse events (AEs) with transcutaneous diaphragmatic stimulation include transient discomfort or pain, muscle fatigue, and skin irritation or burns at the electrode sites; rare cases of arrhythmias have been described. HFT may cause airway dryness or discomfort, oxygen toxicity at high FiO₂, and, in susceptible patients, barotrauma or hemodynamic instability if excessive pressures are generated. However, existing clinical studies in hospitalized, mechanically ventilated, or tracheostomized patients have found these interventions to be generally safe when used with appropriate monitoring and conservative settings [29–31, 32–36].

Combining EDP with HFT may synergistically improve weaning outcomes by simultaneously targeting diaphragmatic function and respiratory load. To date, no randomized controlled trial has evaluated this combination in tracheostomized patients requiring PMV. This study aims to fill this gap by evaluating the efficacy and safety of EDP + HFT compared with standard care alone.

### Objectives {7}

The primary objective is to assess whether EDP combined with HFT reduces the time to successful 24-h separation from invasive mechanical ventilation within 28 days after randomization, compared with standard care alone, in tracheostomized adult patients requiring PMV.

Secondary objectives are to evaluate the effect of EDP + HFT, versus standard care alone, on:

– Changes in diaphragmatic thickness and thickening fraction;
– ICU length of stay;
– 28-day survival;
– Safety, including AEs related to EDP and HFT.

### Trial design {8}

This study is a single-center, randomized, controlled, open-label trial with an allocation ratio of 1:1. The trial tests the hypothesis that the combination of EDP and HFT will improve weaning outcomes compared with standard care alone in tracheostomized patients requiring PMV.

The study protocol follows the Standard Protocol Items: Recommendations for Interventional Trials (SPIRIT) 2013 statement [37]. The study is registered with the Chinese Clinical Trial Registry (ChiCTR2500098295). A SPIRIT schedule summarizing enrollment, interventions, and assessments is provided in **Fig 1**. The flow diagram of the trial is illustrated in **Fig 2**. S1 File shows the SPIRIT checklist. S2 and 3 File show the original protocol approved by the ethics committee.

**Fig 1.**
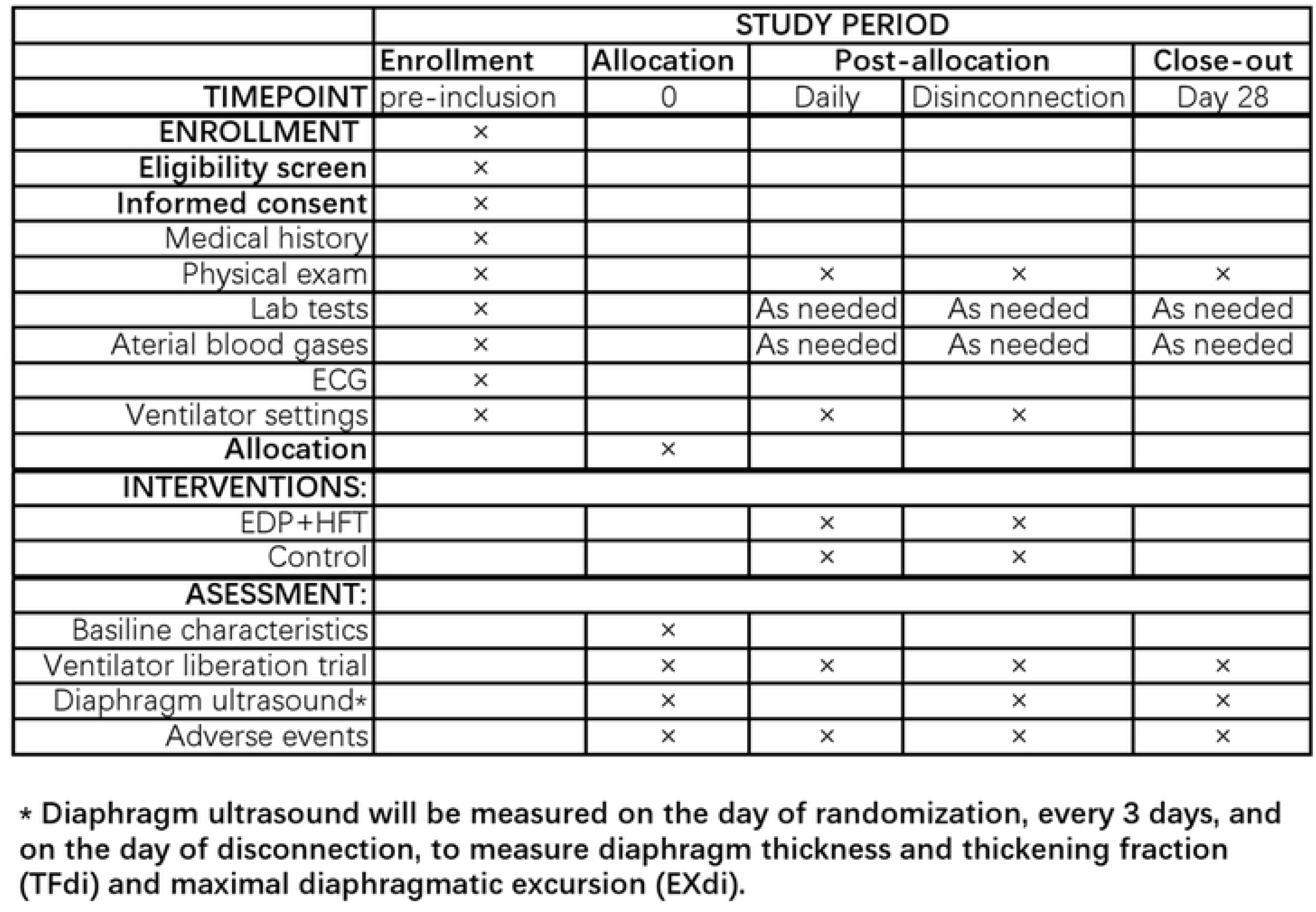
SPIRIT schedule describing study procedures and assessments. SOFA: sequential organ failure assessment, HFT: high-flow oxygen via tracheostomy, EDP: external diaphragm pacing, ECG: electrocardiogram.

**Fig 2.**
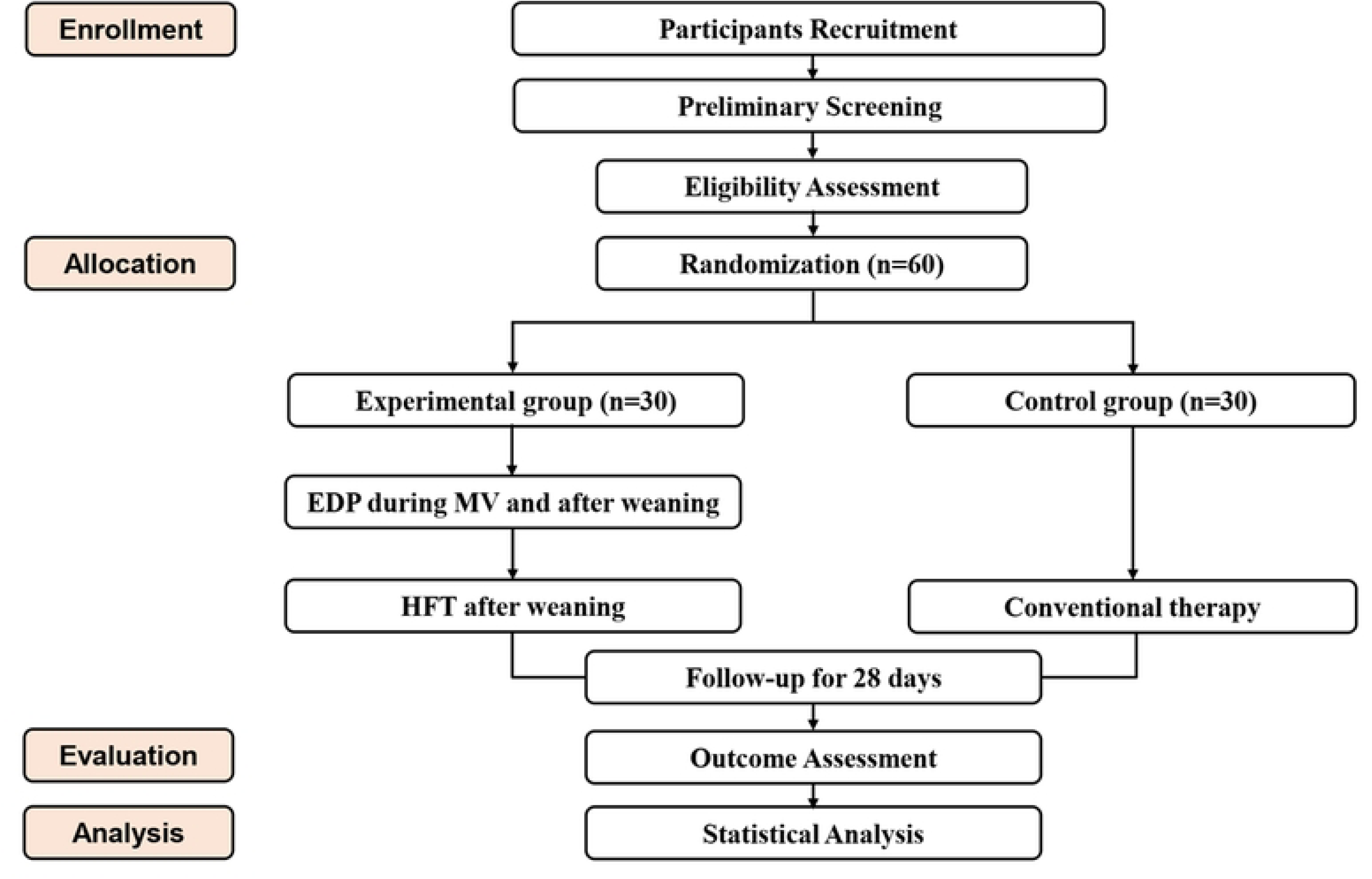
Study flow diagram. EDP: external diaphragm pacing, HFT: high-flow oxygen via tracheostomy, MV: mechanical ventilation.

## Methods: participants, interventions, and outcomes

### Study setting {9}

This study will be conducted at Ningbo Zhenhai Longsai Hospital, a rehabilitation hospital located in Ningbo, Zhejiang Province, China. All study procedures—including participant screening, randomization, delivery of interventions, and outcome assessment—will take place within the ICU of this hospital.

### Eligibility criteria {10}

#### Inclusion criteria

Patients will be included if they meet all of the following criteria:

– Tracheostomized before study screening;
– Receiving invasive mechanical ventilation for >21 consecutive days (≥6 h/day);
– Have failed ≥2 weaning attempts;
– Capable of undergoing EDP with no device-specific contraindications (see Exclusion Criteria for details);
– Patient or legal guardian understands the study and provides written informed consent;
– Hemodynamically stable (systolic blood pressure 90–160 mmHg, heart rate 60–120 bpm, no vasopressor use for ≥24 h);
– Aged ≥18 years.

#### Exclusion criteria

Patients will be excluded if they have any of the following:

– Impaired respiratory center with absent spontaneous breathing (e.g., brainstem injury with absent respiratory drive);
– Rapidly progressive neuromuscular diseases (e.g., amyotrophic lateral sclerosis, Guillain–Barré syndrome);
– Current sepsis/septic shock or hemodynamic instability;
– End-stage chronic diseases with life expectancy <6 months (e.g., stage IV cancer, end-stage heart, liver, or renal failure);
– Presence of implanted cardiac pacemakers or implantable cardioverter-defibrillators (ICDs), due to potential interference with transcutaneous electrical stimulation;
– Morbid obesity or chest wall characteristics that, according to the EDP device manufacturer’s instructions, are likely to prevent effective phrenic nerve stimulation (e.g., BMI ≥35 kg/m² if specified);
– Active infection, burns, open wounds, or significant skin lesions at or near the planned electrode application sites (neck and upper chest);
– Known allergy or intolerance to electrode materials or conductive gels used with the EDP device;
– Pregnancy or lactation.

### Who will take informed consent? {26a}

Before any study-related activities begin, an investigator will provide each patient (or their legally authorized representative, LAR) with details of the study using simple language. The patient will receive a patient information sheet and informed consent form and will have time to review the materials, ask questions, and make an informed decision about voluntary participation. Participation is entirely optional and will not affect standard treatment in any way. Patients may withdraw from the study at any time.

If the patient has decision-making capacity and can sign, they will sign and date the informed consent form in the presence of the investigator, who will also sign. If the patient has capacity but is physically unable to sign (e.g., profound weakness), consent will be obtained verbally or via a clear non-verbal signal in the presence of an impartial witness, while the LAR signs the form on the patient’s behalf. If the patient lacks capacity (e.g., unconsciousness, delirium), consent will be obtained from the LAR. When such patients later regain capacity, the research team will explain the study and seek the patient’s own written consent to continue participation and use of previously collected data.

The patient will receive a copy of the signed form; another copy will be filed with the study documents. Once consent is obtained, the participant will be officially enrolled.

### Additional consent provisions for collection and use of participant data and biological specimens {26b}

Not applicable. No biological samples or additional consent data will be collected.

## Interventions

### Explanation for the choice of comparators {6b}

The control group will receive the current standard of care for PMV weaning, chosen as the comparator for its widespread use in pragmatic trials, its reflection of real-world clinical practice, and its ability to control for care variations over the study period. Patients in this group will not receive EDP or HFT. Instead, they will receive conventional oxygen therapy via a tracheostomy mask, with the fraction of inspired oxygen titrated to maintain peripheral oxygen saturation (SpO₂) ≥92%, consistent with standard oxygenation targets for PMV patients. Except for the experimental group’s use of EDP + HFT, both groups will follow identical study procedures and the same weaning assessment protocol [38, 39], including daily screening for weaning readiness and standardized spontaneous breathing trials (SBTs).

### Intervention description {11a}

#### External diaphragm pacing (EDP)

Patients assigned to the experimental group will receive EDP in addition to standard care. The device used will be the EDP-2023 model manufactured by Arahelio Biotechnology Developing Guangzhou Co., Ltd. (Guangzhou, China). Patients will be positioned supine with the head of the bed elevated to 60 degrees. The upper electrode will be attached to the outer one-third of the lower end of the sternocleidomastoid muscle, while the lower electrode will be placed in the second intercostal space along the mid-clavicular line.

EDP parameters will be set as follows: pulse frequency 40 Hz, stimulation rate 9 breaths per minute, and session duration 30 minutes. Patients will undergo two EDP sessions per day, five days per week, from randomization until the earliest of: (1) successful 24-h ventilator separation; (2) Day 28; (3) ICU discharge; or (4) discontinuation due to AEs or withdrawal of consent. The stimulation intensity will be titrated incrementally from a low level to the maximum tolerated by the patient, with an upper limit of 20 mA to minimize discomfort or pain.

EDP sessions will be delivered by respiratory therapists who have completed standardized training in electrode placement, device operation, safety monitoring, and adverse event reporting. Competency will be documented before independent practice.

#### High-flow oxygen via tracheostomy (HFT)

In addition to EDP, patients in the experimental group will receive HFT using a specialized high-flow system (e.g., Fisher & Paykel Optiflow™) adapted for tracheostomy use. HFT will be initiated during the first SBT or ventilator disconnection period after randomization, delivered through the tracheostomy interface.

Initial parameters for HFT will be: flow rate 60 L/min, gas temperature 37 °C, and fraction of inspired oxygen (FiO₂) 40%. During the weaning period, these parameters will be adjusted dynamically based on arterial blood gas results, circulatory status, and respiratory status, aiming to maintain SpO₂ ≥92% and arterial carbon dioxide partial pressure (PaCO₂) ≤50 mmHg. HFT may be continued as post-weaning support after successful 24-h ventilator separation at the discretion of the treating physician.

#### Standard care (both groups)

All patients will receive standard ICU care for PMV, including:

– Evidence-based weaning assessments and SBTs [38, 39];
– Nutritional support (enteral/parenteral nutrition);
– Antibiotic therapy for infections as indicated;
– Analgesia and sedation management per local protocols;
– Passive and, when appropriate, active limb physical therapy.

### Criteria for discontinuing or modifying allocated interventions {11b}

EDP and/or HFT will be temporarily modified or permanently discontinued if:

– AEs occur that are attributable or possibly attributable to the intervention (e.g., EDP-related skin irritation/burns, intolerable discomfort, suspected arrhythmia; HFT-related oxygen toxicity, barotrauma, severe intolerance). Depending on severity, this may involve reducing EDP intensity, lowering HFT flow or FiO₂, or stopping the intervention.
– Clinical deterioration arises (e.g., severe arrhythmia, acute respiratory failure, hemodynamic instability), requiring the interventions to be paused until stability is restored and the clinical team judges it safe to resume.
– Patient or LAR request for withdrawal from the study, in which case all study-specific interventions will cease and standard care will continue.

All modifications and discontinuations will be documented in the case report forms.

### Strategies to improve adherence to interventions {11c}

Adherence is facilitated by the fact that all interventions occur during a single ICU stay and are administered by trained staff. Investigators will communicate carefully with patients and family members to build trust, clearly explain the importance of the interventions, and ensure understanding before consent. The research team will maintain frequent contact with participants to sustain engagement.

Before enrolling a patient, the team will consider whether any factors (e.g., severe agitation, refusal of procedures) might compromise adherence and will balance the anticipated benefits against such concerns.

### Relevant concomitant care permitted or prohibited during the trial {11d}

#### Permitted concomitant care includes

– Nutritional support (enteral/parenteral);
– Antibiotic therapy and other pharmacological treatments as clinically indicated;
– Analgesia, sedation, and delirium management;
– Passive and active limb physical therapy and mobilization as tolerated.

#### Prohibited care includes

– Other diaphragmatic training techniques (e.g., inspiratory muscle training devices, alternative forms of phrenic nerve stimulation);
– Non-invasive ventilation used specifically as a weaning support during SBTs or post-disconnection periods, to avoid confounding the intervention effect.

### Provisions for post-trial care {30}

Patients in the experimental group who achieve weaning success by day 28 may continue EDP or HFT after trial completion at the clinician’s discretion, but no study funding supports post-trial use of these interventions. No specific insurance or financial compensation is available for trial-related harm. Any intervention-related injuries will be managed according to standard hospital policies and national regulations; costs attributable to trial procedures will be borne by the hospital where applicable.

### Outcomes {12}

#### Primary outcome

– Time (in days) from randomization to successful 24-h separation from invasive mechanical ventilation via the tracheostomy, defined as continuous spontaneous breathing on a tracheostomy collar or HFT without reconnection to the mechanical ventilator within the subsequent 48 h. Decannulation is not required to meet this definition. Patients who die, withdraw life support, or remain ventilator-dependent by day 28 will be treated as having no event and will be included in competing-risk analyses.

#### Secondary outcomes

– Diaphragmatic ultrasound measures: changes in diaphragm thickness and thickening fraction from baseline to each follow-up assessment (every 3 days) and on the day of final ventilator disconnection or Day 28, whichever occurs first.
– Diaphragmatic excursion: maximal diaphragmatic excursion assessed by ultrasound at the same time points.
– ICU length of stay: number of days from ICU admission to ICU discharge.
– 28-day survival: survival status at day 28 after randomization.

#### Safety outcomes

– Incidence and severity of AEs and serious AEs (SAEs) potentially related to EDP and/or HFT, including but not limited to:

* Skin irritation, burns, or pain at electrode sites;
* Arrhythmias, hemodynamic instability;
* Respiratory failure, barotrauma, or oxygen toxicity;
* Other clinically relevant events judged by the investigator to be related or possibly related to the study interventions.

All outcomes are chosen to directly evaluate the efficacy and safety of the combined intervention for weaning in PMV patients.

### Participant timeline {13}

The participant timeline will include the following phases:

#### Screening phase (pre-randomization)

Eligibility assessment based on inclusion and exclusion criteria; informed consent from the patient or LAR; collection of baseline data, including demographics, comorbidities, prior duration of mechanical ventilation and tracheostomy, ventilator settings, arterial blood gas values, and severity scores (e.g., SOFA, APACHE II). Baseline diaphragm ultrasound measurements (thickness, thickening fraction, excursion) and vital signs will be recorded.

#### Randomization (Day 0)

Eligible and consented participants will be randomized 1:1 to the experimental or control group. For participants in the experimental group, EDP and HFT will be initiated as per protocol.

#### Intervention and follow-up period (Day 1–27)

Daily activities will include weaning readiness screening, SBTs, monitoring of vital signs and SpO₂, and adverse event monitoring in both groups. In the experimental group, EDP sessions and HFT will be administered according to the intervention schedule. Diaphragm ultrasound will be performed on the day of randomization (baseline), every 3 days thereafter, and on the day of final ventilator disconnection or Day 28, whichever occurs first. Laboratory tests and arterial blood gases will be conducted as clinically indicated.

#### Final assessment (Day 28 or earlier if early termination)

Final assessment will include weaning status, total time to successful ventilator separation, ICU length of stay, 28-day survival status, and documentation of all AEs/SAEs. Study participation concludes at this point.

### Sample size {14}

This trial involves a novel combined therapy (EDP + HFT) for patients with PMV. As this specific combination has not been previously evaluated in an RCT setting, robust background data to support a conventional power calculation based on precise effect sizes are limited.

The sample size was determined based on clinical relevance and prior experience from similar studies in this population, such as the RESCUE-2 study [28]. We aim to enroll 60 participants (30 per group). While the study is conducted at a single center, this sample size is estimated to be sufficient to detect clinically meaningful trends in the primary endpoint and provide a comprehensive safety profile. All randomized participants will be included in the primary intention-to-treat analysis.

### Recruitment {15}

Eligible tracheostomized PMV patients will be identified through daily screening by the study nurse and referrals from ICU attending physicians. The research team will inform patients and/or guardians about the trial’s objectives, procedures, potential benefits, and risks to facilitate informed decision-making and support recruitment. Recruitment progress will be tracked using a weekly dashboard reviewed by the Trial Management Group.

## Assignment of interventions: allocation

### Sequence generation {16a}

Permuted block randomization will be used, with variable block sizes of 4 and 6 to prevent predictability of group allocation. A computer-generated randomization sequence will be created using SAS 9.4 software by an independent biostatistician not involved in patient care.

### Concealment mechanism {16b}

The randomization sequence will be stored in sequentially numbered, opaque, sealed envelopes. Each envelope will contain the treatment allocation for a unique study ID. Envelopes will be opened only after the patient’s eligibility has been confirmed and informed consent obtained.

### Implementation {16c}

Participants will be enrolled by the research team. The independent biostatistician will generate the allocation sequence and prepare the sealed envelopes. Upon enrollment, the investigator will open the next envelope in sequence to assign the participant to the experimental or control group.

## Assignment of interventions: blinding

### Who will be blinded {17a}

This is an open-label trial: neither participants nor treating clinicians will be blinded to treatment arm. However, to reduce detection and assessment bias:

– Sonographers performing diaphragm ultrasound measurements will be blinded to treatment allocation whenever feasible and will follow standardized protocols;
– Stored ultrasound images will be anonymized and a subset will be reviewed centrally by a blinded expert for quality control;
– Statisticians performing the primary and secondary analyses will be blinded to group allocation.

The primary endpoint—time to successful 24-h ventilator separation—is an objective measure derived from ventilator logs and medical records by staff who are not involved in delivering the interventions.

### Procedure for unblinding if needed {17b}

The study is unblinded for participants and treating clinicians throughout. Blinding of ultrasound assessors and statisticians will not normally be broken. In exceptional circumstances where knowledge of allocation is deemed essential for safety or data interpretation, the principal investigator may authorize unblinding of the relevant assessor; such instances will be documented.

## Data collection and management

### Plans for assessment and collection of outcomes {18a}

All assessments will be conducted by qualified professionals delegated by the principal investigator.

#### Baseline data

At screening and prior to randomization, baseline data will be collected from medical records and clinical assessments, including:

– Demographics (age, sex, BMI);
– Primary diagnosis leading to respiratory failure;
– Major comorbidities;
– Duration of mechanical ventilation and tracheostomy at randomization;
– Ventilator settings and arterial blood gas values;
– Severity of illness scores (e.g., SOFA, APACHE II);
– Diaphragm ultrasound measurements (thickness, thickening fraction, excursion).

#### Diaphragm ultrasound

Diaphragmatic thickness and thickening fraction will be measured using B-mode ultrasound in the zone of apposition of the right hemidiaphragm. A linear or curvilinear probe will be placed at the 8th–10th intercostal space between the anterior and mid-axillary lines. Three consecutive respiratory cycles during tidal breathing will be recorded, and measurements will be averaged across cycles. Maximal diaphragmatic excursion will be assessed using M-mode ultrasound with the probe placed subcostally.

Sonographers will complete a training program and demonstrate acceptable inter- and intra-observer variability before participating. Ultrasound images will be stored digitally; a random 10% sample will be re-evaluated by a blinded expert for quality control.

#### Daily and follow-up assessments

From Day 1 to Day 27, daily assessments will include:

– Weaning readiness screening and performance of SBTs per protocol;
– Vital signs and SpO₂;
– Documentation of ventilator settings and any ventilator disconnections;
– Recording of AEs and SAEs.

Diaphragm ultrasound will be repeated every 3 days and on the day of final ventilator disconnection or Day 28, whichever occurs first. ICU length of stay, time to 24-h ventilator separation, and 28-day survival will be recorded.

### Plans to promote participant retention and complete follow-up {18b}

Because the trial interventions and the primary endpoint are confined to the inpatient setting over a short time frame (28 days), loss to follow-up is expected to be minimal. If participants deviate from the protocol or withdraw from the study interventions, outcome data will continue to be collected according to the protocol unless the participant or LAR explicitly withdraws consent for further data use.

Participants discharged from the ICU before Day 28 will be followed until Day 28 or in-hospital death through ward and rehabilitation unit records.

### Data management {19}

Patient information prior to ICU admission will be obtained from the hospital’s electronic medical records. Clinical details and outcomes during the ICU stay will be documented by the intensivists and nursing staff responsible for care and then transferred to study-specific case report forms.

Anonymized data will be entered into an electronic data capture system with built-in range and consistency checks. Only authorized personnel (investigators, data manager, monitor) will have password-protected access. The ethics committee and study monitor will oversee data collection and management processes to ensure accuracy, completeness, and protocol compliance.

### Confidentiality {27}

Participants will be identified in the electronic case report form (eCRF) only by a unique study ID. A linkage file connecting names to study IDs will be stored in a locked cabinet at the study site, accessible only to investigators and delegated staff. Electronic data will be stored on secure servers with regular backups. All data handling will comply with Chinese privacy regulations, and access will be limited to authorized personnel.

### Plans for collection, laboratory evaluation, and storage of biological specimens for genetic or molecular analysis in this trial/future use {33}

Not applicable. This trial will not involve collection, laboratory evaluation, or storage of biological specimens for genetic or molecular analysis, either for the current study or for future research.

## Statistical methods

### Statistical methods for primary and secondary outcomes {20a}

Analyses will be conducted on two populations:

– Intention-to-treat (ITT) population: all randomized participants, analyzed according to their assigned group;
– Per-protocol (PP) population: participants who complete the study without major protocol deviations.

#### Primary outcome

Time to successful 24-h ventilator separation will be analyzed using Kaplan–Meier survival curves and log-rank tests. Death and withdrawal of life support before weaning will be treated as competing risks; cumulative incidence functions and Gray’s test will be used for sensitivity analyses. Hazard ratios and 95% confidence intervals will be estimated using Cox proportional hazards models, if appropriate.

#### Secondary outcomes

– Changes in diaphragmatic thickness and thickening fraction over time will be analyzed with mixed-effects linear models including fixed effects for treatment group, time, and group × time interaction, with random intercepts for participants.
– ICU length of stay, which is often skewed, will be compared between groups using non-parametric tests (e.g., Mann–Whitney U test) and summarized using medians and interquartile ranges.
– 28-day survival and incidence of AEs/SAEs will be compared using Fisher’s exact test or chi-square test as appropriate.

Analyses will be performed using standard statistical software (e.g., SAS, R). All tests will be two-sided with a significance level of 0.05.

### Methods for additional analyses (e.g., subgroup analyses) {20b}

Given the sample size, no formal subgroup or multiple sensitivity analyses are planned. Post-hoc analyses may examine associations between baseline characteristics (e.g., diaphragm thickness, duration of mechanical ventilation before randomization) and outcomes.

### Methods in analysis to handle protocol non-adherence and any statistical methods to handle missing data {20c}

Protocol deviations and non-adherence (e.g., missed EDP sessions) will be documented and summarized. The primary analysis will use the ITT population; the PP analysis will provide supportive information.

For longitudinal outcomes (e.g., diaphragm ultrasound measures), mixed-effects models will use maximum likelihood estimation, which accommodates missing measurements under a missing at random (MAR) assumption without requiring explicit imputation. For time-to-event outcomes, standard survival and competing-risk methods will censor participants at their last known follow-up. Given the short, inpatient follow-up window, the extent of missing data is expected to be limited. No single-value imputation methods (such as last observation carried forward) will be used.

### Interim analyses {21b}

No interim efficacy or futility analyses are planned due to the relatively small sample size and short duration of the intervention.

### Plans to give access to the full protocol, participant-level data, and statistical code {31c}

The full study protocol is available from the corresponding author upon reasonable request. Before publication, access to participant-level data will be restricted to the research team. After publication, de-identified participant-level data and statistical code will be shared via a public data repository in accordance with a data-sharing agreement and applicable regulations.

## Oversight and monitoring

### Composition of the coordinating centre and trial steering committee {5d}

This single-center study does not have a formal coordinating center or external trial steering committee. Overall trial conduct is overseen by the Trial Management Group as described above.

### Composition of the data monitoring committee, its role, and reporting structure {21a}

A study monitor, independent of the clinical care team and not involved in patient recruitment, intervention delivery, or data analysis, will be responsible for routine data monitoring, auditing, and reviewing adverse event documentation. The monitor will perform site visits at trial initiation, during recruitment, and at study close-out to:

– Check completeness and accuracy of source documents and eCRFs;
– Verify data consistency and resolve discrepancies;
– Ensure adherence to the protocol, GCP, and informed consent procedures.

In addition, an Independent Safety Officer will periodically review aggregated safety data, focusing on SAEs and device-related complications. The Safety Officer may recommend protocol modifications or early termination if safety concerns arise. Recommendations will be communicated to the principal investigator and the ethics committee.

### Adverse event reporting and harms {22}

An AE is defined as any untoward medical occurrence in a participant administered a trial intervention, whether or not it is considered related to that intervention. A SAE is an AE that results in death, is life-threatening, requires or prolongs hospitalization, results in persistent or significant disability/incapacity, or is an important medical event according to the investigator’s judgment.

All AEs will be recorded in the eCRF with details on onset, duration, severity, outcome, and relationship to the study interventions. SAEs judged to be related or possibly related to EDP or HFT, or that are unexpected, will be reported to the ethics committee and the Independent Safety Officer within 24 hours of the investigator becoming aware of the event, in accordance with ICH-GCP and local regulatory requirements.

### Frequency and plans for auditing trial conduct {23}

The hospital’s Department of Medical Education, on behalf of the sponsor, will audit the trial approximately every 6 months to assess:

– Completeness and accuracy of source documents and case report forms;
– Protocol compliance and adherence to ethical standards;
– Appropriate reporting and handling of AEs and SAEs.

The funding body may conduct one external audit during the recruitment period to evaluate data integrity and appropriate use of funds.

### Plans for communicating important protocol amendments to relevant parties (e.g., trial participants, ethical committees) {25}

Any substantial amendments to the study protocol will require prior approval from the ethics committee. If changes affect participant information or consent, the informed consent documents will be revised and re-consent will be obtained as necessary. Protocol amendments will be documented and, where relevant, updated in the trial registry and reported in subsequent publications.

### Dissemination plans {31a}

Trial results will be submitted to a peer-reviewed journal within one year of trial completion. Reporting will follow CONSORT guidelines for randomized trials. Key findings will also be disseminated through oral and poster presentations at national and international critical care and respiratory conferences. For broader accessibility, a plain-language summary of the results will be provided to trial participants, ICU staff, and the public via hospital newsletters and official social media channels.

## Trial status

The current protocol (version 1.1) was approved on 1 March 2025. Recruitment is planned to start on 1 November 2025 and to conclude by 30 April 2026. Data collection will be completed by 31 August, 2026, with the trial results anticipated to be available by 31 December, 2026.

## Discussion

This randomized controlled trial addresses an important clinical gap by evaluating the synergistic effect of combining external diaphragm pacing with high-flow oxygen via tracheostomy to facilitate weaning in patients requiring prolonged mechanical ventilation. This population is characterized by high morbidity, VIDD, and resource utilization. EDP targets diaphragm activation and may counteract VIDD, while HFT stabilizes oxygenation, reduces work of breathing, and is generally well tolerated in tracheostomized patients.

We chose HFT rather than non-invasive ventilation for three main reasons: (1) tracheostomized patients often have reduced upper airway resistance, making HFT-mediated dead space reduction more efficient; (2) HFT delivered via tracheostomy is typically better tolerated than tight-fitting masks required for non-invasive ventilation, thereby minimizing discomfort during weaning; and (3) existing data in tracheostomized populations indicate that HFT can improve oxygenation and reduce re-intubation risk while avoiding some barotrauma risks associated with non-invasive ventilation [33, 35, 36].

Key strengths of this study include being, to our knowledge, the first RCT of this non-invasive combination in PMV patients, adherence to SPIRIT guidelines, and use of objective, clinically relevant outcomes with blinded assessment wherever feasible. The trial also incorporates rigorous diaphragm ultrasound measurements and standardized weaning protocols.

However, several limitations must be acknowledged. The study is single-center and open-label, which may limit generalizability and introduce performance bias, although the primary outcome is objective. The sample size is relatively small, meaning the trial may have limited power to detect smaller effect sizes.

If EDP + HFT is shown to be safe and associated with shorter time to ventilator liberation and improved diaphragm function, the findings will provide evidence to support this strategy and justify future multicenter trials.

## Abbreviations

eCRF: Electronic case report form
EDP: External diaphragm pacing
HFT: High-flow oxygen via tracheostomy
ICU: Intensive care unit
LAR: Legally authorized representative
MAR: Missing at random
PMV: Prolonged mechanical ventilation
RCT: Randomized controlled trial
SAE: Serious adverse event
SBT: Spontaneous breathing trial
SOFA: Sequential Organ Failure Assessment
VIDD: Ventilator-induced diaphragmatic dysfunction

## Authors’ contributions {31b}

Lei Yang conceptualized and drafted the protocol. Kebin Zheng, Na Li, Kongting Xing, and Jing Zhang contributed to study design, critical revision of the manuscript, and approved the final version. All authors agree to be accountable for all aspects of the work.

## Acknowledgements

We thank Dr. Federico Longhini (Magna Graecia University of Catanzaro, Italy) and Dr. Gerrard Rafferty (King’s College London, UK) for their critical review of the protocol. We also thank the nursing staff of Ningbo Zhenhai Longsai Hospital ICU for their support in recruitment and data collection.

## Data availability {29}

Before publication, access to the full dataset will be restricted to the research team. After publication, de-identified participant-level data and statistical code will be made available from the corresponding author on reasonable request and will be shared via a public repository, in line with hospital policies and applicable regulations.

## Declarations

### Ethics approval and consent to participate {24}

The study was approved by the Medical Ethics Committee of Ningbo Zhenhai Longsai Hospital (Approval No. 2023-003, dated 29 December 2023). Written informed consent will be obtained from all participants or their legal guardians before enrollment.

### Consent for publication {32}

Not applicable. No individual patient data or images will be included in publications.

### Supporting Information

None

